# Longitudinal Associations between Hair Cortisol, PTSD Symptoms, and Sleep Disturbances in a Sample of Firefighters with Duty-related Trauma Exposure

**DOI:** 10.1101/2021.08.22.21262402

**Authors:** M. R. Sopp, T. Michael, J. Lass-Hennemann, S. Haim-Nachum, M. J. J. Lommen

## Abstract

Several studies have found evidence of altered cortisol levels in patients with posttraumatic stress disorder (PTSD). Based on these findings, it is assumed that these patients may show signs of cortisol dysregulation after trauma. Posttrauma cortisol levels are thus considered a potential biomarker of PTSD. However, longitudinal studies using indicators of long-term cortisol secretion (such as hair cortisol concentrations; HCC) are scarce. The current study investigated prospective associations between HCC and PTSD symptoms in a sample of Dutch firefighters taking into account varying levels of work-related trauma severity. In addition, we assessed posttraumatic sleep disturbances as a secondary outcome measure to investigate whether effects generalize to this frequent comorbidity of PTSD. Three hundred seventy-one Dutch firefighters with a mean of 14.01 years of work experience were included in the analyses. Baseline assessment included the collection of hair samples and the measurement of work-related trauma severity, PTSD symptoms, and sleep disturbances. PTSD symptoms and sleep disturbance were re-assessed after six and twelve months. Multilevel analyses indicate a significant positive correlation between HCC and baseline PTSD symptoms in those with average or above-average work-related trauma severity. A similar pattern was evident for posttraumatic sleep disturbances at baseline. Moreover, higher HCC predicted more posttraumatic sleep disturbances after 6 months in participants with above-average work-related trauma severity. No other associations emerged for PTSD symptoms or posttraumatic sleep disturbances at six or twelve months. As such, our study supports the existence of a cross-sectional association between HCC and trauma symptoms, which may vary for different levels of subjective trauma severity. The longitudinal stability of this association should be reinvestigated by future research.

---

The majority of the current world population will experience trauma during their lifetime (APA, 2013; Kessler et al., 2017). However, some individuals – such as firefighters (Jahnke et al., 2016) – are at a heightened risk (Lewis-Schroeder et al., 2018). Critically, firefighters are not only exposed to occupational trauma once but repeatedly, which is assumed to increase their risk of developing mental disorders (Geronazzo-Alman et al., 2017). One of the mental disorders that may arise from occupational trauma is posttraumatic stress disorder (PTSD). PTSD prevalence in first responders differs considerably across studies and may vary as widely as from 0 to 46% (Soravia et al., 2021). Importantly, PTSD is not the only potential sequel of occupational trauma. Besides others (e.g., depression and substance use disorders; Kim et al., 2018), sleep disturbances are a common trauma-related mental health problem (Werner et al., 2021) seen in up to 69% of firefighters (Abbasi et al., 2018; Carey et al., 2011).

Despite these detrimental effects of occupational trauma, firefighters experience fewer posttrauma symptoms on average than individuals from the general population involved in the same incidents (Gulliver et al., 2021; Schnell et al., 2020). These effects may emerge because only individuals with high trait resilience choose this profession or due to specific characteristics of occupational trauma (e.g., psychological preparedness). However, although the majority of firefighters may be resilient, some do experience symptoms, warranting further research into the pathogenic processes underlying mental health problems following occupational trauma (Schnell et al., 2020). With respect to PTSD, current research points towards a critical role of endocrinological stress responses during and after trauma (van Zuiden et al., 2019). Acute stress activates the hypothalamic-pituitary-adrenal (HPA) axis, resulting in the secretion of corticotropin-releasing hormone (CRH) in the thalamus. CRH stimulates the release of adrenocorticotropic hormone in the pituitary gland, which, in turn, causes the adrenal cortex to produce cortisol (Pan et al., 2020; Pan et al., 2018). This stress-induced secretion of cortisol has several metabolic effects, allowing the individual to retain more energy that can be mobilized to cope with the stressor. At the same time, it initiates the downregulation of subsequent cortisol levels, restoring the body back to its pre-stressor state. The stress response of the HPA-axis thus allows optimal adaption to short-term stressors, whereas long-term stress may result in dysregulation of cortisol secretion.

In light of these patterns, research has investigated cortisol dysregulation after trauma as a potential precursor of PTSD. Although associations are found in numerous studies, the direction of this association is inconsistent with some studies finding a positive and others a negative link between basal cortisol levels and PTSD symptoms (Meewisse et al., 2007; Schumacher et al., 2019). When looking for sources of these discrepancies, it is important to note that most studies used saliva samples to assess cortisol. Although saliva cortisol is reflective of long-term cortisol dysregulation, it is also susceptible to short-term changes in cortisol secretion (Stalder & Kirschbaum, 2012). Thus, recent research has focused on assessment methods that are better suited for capturing long-term cortisol dysregulation, the most prominent approach being hair cortisol concentrations (HCC).

Based on recent studies assessing HCC (e.g., Luo et al., 2012; Steudte-Schmiedgen et al., 2015; Steudte et al., 2011), Steudte-Schmiedgen et al. (2016) propose that early after trauma, patients with PTSD may show elevated cortisol levels. In the long-term, however, they may show a chronic attenuation of cortisol levels, which is strengthened by subsequent trauma. This model is consistent with the putative effects of long-term stress on the negative feedback loop involved in cortisol secretion. However, in contrast to this model, some studies (Buchmüller et al., 2020; Pacella et al., 2017) have found higher HCC in patients with PTSD as compared to traumatized controls, even after temporally distant traumatization. That is, van Heuvel et al. (2020) recently found higher HCC in adult patients with PTSD after multiple trauma exposure related mostly to childhood maltreatment. The authors interpret these findings as indicating that enhanced – rather than reduced – HCC may emerge in high-trauma environments and in settings with ongoing stressors. Correspondingly, Steudte-Schmiedgen et al. (2016) discuss potential modulatory effects of trauma characteristics (e.g., trauma severity, ongoing vs. distinct traumatization) and comorbidities (e.g., comorbid depression and/or anxiety symptoms) on the relationship between HCC and PTSD symptoms. Further characterizing these effects requires gathering data from different trauma settings (Buchmüller et al., 2020).

Critically, there is a dearth of studies investigating longitudinal associations between HCC and PTSD symptoms, which is vital to test the model proposed by Steudte-Schmiedgen et al. (2016). To our knowledge, four studies have investigated longitudinal associations between HCC and PTSD symptoms. Of these studies, one study found a negative association between posttrauma HCC and subsequent PTSD symptoms (Steudte-Schmiedgen et al., 2015), whereas one found a positive association (Pacella et al., 2017), and two did not find a significant association (Petrowski et al., 2020; Straub et al., 2017). However, sample sizes varied considerably between studies (*N* = 30 - 90) with only one study being sufficiently powered to detect medium-sized correlations. Moreover, no study to date has investigated the temporal stability of the association between HCC and PTSD symptoms and the extent to which it is modulated by trauma severity.

In addition, few studies have investigated associations between HCC and other mental health problems that may result from trauma. Specifically, associations between HCC and sleep disturbances remain yet to be investigated. These associations are of specific interest due to the strong bidirectional links between cortisol levels and sleep physiology (Born et al, 1988; Rodenbeck et al., 2002). Correspondingly, studies in non-trauma-related occupational settings demonstrate that higher HCC is linked with greater sleep disturbances (Wang et al., 2019; Zhang et al., 2020). Moreover, research shows that posttraumatic sleep disturbances are one of the most frequent comorbidities of PTSD (70-91%; Colvonen et al., 2018), while simultaneously indicating that they are not just a symptom of PTSD. They rather seem to constitute an independent sequel of trauma with bidirectional links to PTSD (Roberge & Bryan, 2021).

To address the gaps outlined above, we conducted a study investigating longitudinal associations between HCC (assessed at baseline) and PTSD symptoms (assessed at baseline, + six months, and + twelve months) in a comprehensive sample (*N* = 371) of firefighters with repeated trauma exposure. To account for differential effects of trauma exposure, we investigated moderating effects of work-related trauma severity (WRTS) assessed at baseline on the association between HCC and PTSD symptoms. As an additional outcome measure, we assessed sleep disturbances. Previous research (Steudte-Schmiedgen et al., 2016) indicates that patients with PTSD show elevated cortisol levels early after trauma and a chronic attenuation after long delays and subsequent trauma. Since our sample had repeatedly experienced duty-related trauma at the time of baseline assessment, we hypothesized that correlational patterns would reflect chronic attenuation of cortisol levels. As such, we hypothesized to find a negative association between HCC and PTSD symptoms. Moreover, we expected this association to increase with time and subsequent trauma exposure. Finally, we hypothesized to find a positive correlation between HCC and sleep quality, which we also expected to increase with time.

## Materials and Methods

### Participants

Five hundred twenty-nine participants employed as firefighter in the Netherlands (The Netherlands Fire Service) enrolled in this study (for other publications on this sample see Egberink et al., 2020 and de Haart et al., 2021). Of these, 157 participants could not be included in hair analyses because they either failed to provide scalp hair of sufficient length or opted not to provide a hair sample. Of all those providing hair samples, one participant was excluded because the determined HCC was higher than five standard deviations above the mean. Thus, the final sample comprised 371 participants (92.72% male; *M*_age_= 38.78, *SD*_age_= 10.10) at baseline. In this still ongoing study, follow-up assessments are administered every half year. Due to the limited sample of participants providing valid hair samples and high participant loss across time, we decided to restrict our analyses to the first and second follow-up assessments. For the PTSD measure, response rates were 64.15% (*n* =238) at the first follow-up (six months) and 47.17% (*n* =175) at twelve months. The study protocol was approved by the ethics committee of Behavioural and Social Sciences at the University of Groningen and all participants gave written informed consent in accordance with the Declaration of Helsinki.

The study was conducted as part of a larger project that aimed to recruit as many firefighters as possible from 25 safety regions in the Netherlands. Post-hoc power analyses (*α* = .05 - two-tailed, 1-*β* = .85) indicate that our sample size was sufficiently large to detect small-to-medium correlations (Baseline: *r* = .15, 6 months: *r* = .19, 12 months: *r* = .22). As such, the current study was sufficiently powered to detect associations found by prior research (Pacella et al., 2017; Steudte-Schmiedgen et al., 2015).

### Assessment of Self-Report Measures

Eleven out of 25 safety regions in the Netherlands agreed to participate in this study. Written information about the study was sent to representatives of the participating regions, after which a planning was made based on availability of the participating regions during the summer of 2017. Firefighters were approached at individual fire stations, were given oral and written information about the study, and were asked to participate. After providing written informed consent, participants were asked to complete several computer tasks and fill out a battery of self-report questionnaires. Of these questionnaires, we used measures of WRTS, lifetime trauma exposure, psychopathology, PTSD symptoms, and sleep disturbances for the current analyses. Participants were contacted again via email six and twelve months after baseline assessment to assess PTSD symptoms and sleep disturbances in the preceding six-month period through an online questionnaire.

#### WRTS

To assess WRTS, a questionnaire was developed in collaboration with a group of firefighters (see Supplementary Table 1 for details). The questionnaire contains 21 stressful events such as “resuscitation of a child” or “deceased adult by fire”. For each event, participants indicate whether they have experienced the event (yes/no). Moreover, they rate the emotional impact the respective event has had on them (1 = “No emotional impact”, 2 = “Little emotional impact”, 3 = “Quite an emotional impact”, 4 = “High emotional impact”). For the current study, responses were recoded into one scale ranging from 0 to 4 (0 = “I have never experienced this event”, 1 = “No emotional impact”, 2 = “Little emotional impact”, 3 = “Quite an emotional impact”, 4 = “High emotional impact”). The sum score across all recoded items was used for further analyses. Since the questionnaire was developed specifically for this study, validity and reliability have not been investigated yet. However, internal consistency was found to be high (*α* = .871) in the current sample.

#### Lifetime Trauma Exposure

To control for the effects of lifetime trauma exposure, we used the Life Events Checklist for DSM-5 (LEC-5; Boeschoten et al., 2014). The self-report questionnaire contains sixteen potentially traumatic events. Participants are asked to indicate their level of exposure. In accordance with criterion A of the DSM-5 PTSD criteria (APA, 2013), we calculated the sum of events for which participants indicated direct (happened to me) or indirect (witnessed it or part of my job) exposure as our indicator of lifetime trauma exposure.

#### Psychopathology

General psychological symptom burden was assessed using the Dutch version of the Brief Symptom Inventory (BSI; Derogatis & Melisaratos, 1983). The BSI is a 53-item self-report instrument that measures symptomatic distress using nine subscales. The inventory has demonstrated good convergent and construct validity. For this study, the Global Severity Index (GSI), which indicates general psychopathological symptom burden was used for all analyses.

#### PTSD Symptoms

The PTSD Checklist for DSM-5 (PCL-5) was used to assess PTSD symptoms during the past month in accordance with DSM-5 symptom criteria (Boeschoten et al., 2014). The questionnaire comprises 20 items, which are rated on a scale ranging from 0 = “not at all” to 4 = “extremely”. Reliability and validity of the original measure are good (Wortmann et al., 2016). The total score was used for the current analyses.

#### Sleep Disturbances

The Pittsburgh Sleep Quality was used to assess sleep disturbances in the past month (Buysse et al., 1991). The questionnaire consists of 19 items measuring subjective sleep features (e.g., sleep onset latency) and occurrences that might compromise sleep quality (e.g., nightmares). Based on participants’ responses, a score ranging from 0 to 3 is assigned to seven components. Strong reliability and validity, and moderate structural validity of the PSQI have been demonstrated across various translations (Mollayeva et al., 2016). The total score was used for the current analyses.

### HCC Analysis

If participants agreed to take part in HCC assessment and could provide hair of sufficient length, a hair sample was cut from the posterior vertex scalp. The samples were placed into aluminum foil, sealed, and stored in dark containers at room temperature. Analyses were performed at the Biochemical Laboratory of the Department of Biological and Clinical Psychology at University of Trier. The proximal 2 cm of the hair segments were used for analysis, reflecting cortisol secretion for the previous two months based on an accepted growth rate of 1 cm per month (Stalder et al., 2017).

### Data Analyses

A series of multilevel models was fit separately for PTSD symptoms and sleep disturbances. The models included the Level 2 (L2) predictors HCC and WRTS, the Level 1 (L1) predictor Time, and their respective interactions. To account for further sources of variance, the L2 predictors Sex, Type of service (volunteer firefighters vs. professional firefighters with or without voluntary service), Service years, Psychopathology, and Lifetime trauma exposure were introduced as covariates. Aside from dichotomous variables, all L2 variables were grand-mean centered (Kreft et al., 1995). The L1 variable Time was centered at baseline with main effects reflecting the initial status of participants at the beginning of the study (Singer & Willett, 2005).

In a first step, we constructed a baseline model comprising all covariates, WRTS, Time, and the interaction between Time and WRTS as fixed effects (Model 1). Then, we tested the improvement of model fit after inclusion of a random intercept (Model 2) and random slope (Model 3). Finally, we included HCC and the respective interactions with WRTS and Time as fixed effects (Model 4) and assessed the improvement of model fit. All multilevel models were fit using maximum likelihood estimation and the nlme package (Pinheiro et al., 2017) in R (R Core Team, 2013). Significant interactions were probed using simple slopes techniques (Hughes, 2017). Simple slopes were estimated for associations between baseline HCC and outcomes at the mean of WRTS +/-1 *SD*.

## Results

### Sample Characteristics and Bivariate Correlations

Participants worked as volunteer firefighters (29.65%) or professional firefighters with or without voluntary service (70.35%). At the time of baseline assessment, they reported having on average 14.01 years (*SD* = 9.24) of work experience and indicated that they had faced 10.08 (*SD* = 4.24) types of potentially traumatic events during service. The mean HCC was 4.26 pg/mg (*SD* = 4.21; for details on the distribution see Supplementary Table 2).

Bivariate Pearson correlations of all study variables are reported in Table 1. As expected, service years, lifetime trauma exposure, and WRTS were significantly correlated. No consistent correlations emerged between HCC and PTSD symptoms or sleep disturbances. WRTS was consistently correlated with PTSD symptoms while Psychopathology at baseline was consistently correlated with PTSD symptoms and sleep disturbances.

**Table 1.** Bivariate associations between Service years, Lifetime trauma exposure (TE), Psychopathology, Work-related trauma severity (WRTS), Hair cortisol concentration (HCC), and symptoms

| Measures | 1 | 2 | 3 | 4 | 5 | 6 | 7 | 8 | 9 | 10 | 11 |
| --- | --- | --- | --- | --- | --- | --- | --- | --- | --- | --- | --- |
| 1. Service years | 1 | .225** | -.035 | .414** | -.028 | .010 | -.077 | .039 | -.071 | -.103 | -.037 |
| 2. Lifetime TE | .225** | 1 | .088 | .496** | -.025 | .115* | .066 | .166* | .133* | .103 | .097 |
| 3. Psychopathology | -.035 | .088 | 1 | .229** | .030 | .728** | .253** | .291** | .567** | .384** | .372** |
| 4. WRTS | .414** | .496** | .229** | 1 | -.030 | .329** | .139* | .319** | .175** | .023 | .122 |
| 5. HCC | -.028 | -.025 | .030 | -.030 | 1 | .066 | .064 | -.053 | .066 | .145* | -.029 |
| 6. PCL at T0 | .010 | .115* | .728** | .329** | .066 | 1 | .427** | .522** | .522** | .360** | .365** |
| 7. PCL at T1 | -.077 | .066 | .253** | .139* | .064 | .427** | 1 | .549** | .135* | .430** | .347** |
| 8. PCL at T2 | .039 | .166* | .291** | .319** | -.053 | .522** | .549** | 1 | .148 | .268** | .409** |
| 9. PSQI at T0 | -.071 | .133* | .567** | .175** | .066 | .522** | .135* | .148 | 1 | .529** | .635** |
| 10. PSQI at T1 | -.103 | .103 | .384** | .023 | .145* | .360** | .430** | .268** | .529** | 1 | .672** |
| 11. PSQI at T2 | -.037 | .097 | .372** | .122 | -.029 | .365** | .347** | .409** | .635** | .672** | 1 |

*Note.* PCL = PTSD checklist for DSM-5, PSQI = Pittsburgh Sleep Quality Index.
\* $p < .05$ . \*\* $p < .01$

### Prediction of PTSD Symptoms Based on HCC

Repeated assessments of PTSD symptoms were non-independent as reflected in an ICC of .476. The model including a random intercept fitted our data significantly better than the baseline model (χ^2^_diff_ (1) = 50.25, *p* < .0001). Moreover, including a random slope further improved model fit (χ^2^_diff_ (2) = 18.99, *p* = .0001). Finally, including baseline HCC as predictor resulted in further improvements of model fit (χ^2^_diff_ (4) = 9.62, *p* = .047). Table 2 provides an overview of intercepts and slopes as well as the estimated variance accounted for by each model.

**Table 2.** Model summaries of linear mixed model analyses for PTSD symptoms

| *Predictors* | Baseline model |  |  | Random intercept |  |  | Random intercept + Slope |  |  | Random intercept + Slope and<br>Cortisol as predictor |  |  |
| --- | --- | --- | --- | --- | --- | --- | --- | --- | --- | --- | --- | --- |
|  | *Estimates* | *CI* | *p* | *Estimates* | *CI* | *p* | *Estimates* | *CI* | *p* | *Estimates* | *CI* | *p* |
| (Intercept) | 5.37 | 4.45 – 6.30 | **<0.001** | 5.48 | 4.45 – 6.52 | **<0.001** | 5.57 | 4.58 – 6.55 | **<0.001** | 5.61 | 4.62 – 6.60 | **<0.001** |
| Lifetime TE | -0.10 | -0.29 – 0.09 | 0.293 | -0.11 | -0.33 – 0.11 | 0.312 | -0.13 | -0.34 – 0.08 | 0.238 | -0.14 | -0.35 – 0.08 | 0.213 |
| Service years | -0.05 | -0.10 – -0.00 | **0.043** | -0.05 | -0.11 – 0.01 | 0.115 | -0.05 | -0.10 – 0.01 | 0.123 | -0.05 | -0.11 – 0.01 | 0.108 |
| Type of service | -0.56 | -1.05 – -0.07 | **0.025** | -0.54 | -1.11 – 0.03 | 0.067 | -0.39 | -0.95 – 0.16 | 0.166 | -0.38 | -0.94 – 0.18 | 0.186 |
| Psychopathology | 14.67 | 12.79 – 16.55 | **<0.001** | 15.17 | 13.01 – 17.33 | **<0.001** | 16.89 | 14.81 – 18.98 | **<0.001** | 16.57 | 14.46 – 18.68 | **<0.001** |
| Sex | -0.20 | -1.02 – 0.62 | 0.637 | -0.29 | -1.25 – 0.68 | 0.562 | -0.30 | -1.24 – 0.64 | 0.529 | -0.31 | -1.25 – 0.64 | 0.526 |
| Time | -0.17 | -0.25 – -0.08 | **<0.001** | -0.18 | -0.25 – -0.10 | **<0.001** | -0.18 | -0.27 – -0.10 | **<0.001** | -0.19 | -0.28 – -0.11 | **<0.001** |
| WRTS | 0.13 | 0.07 – 0.18 | **<0.001** | 0.13 | 0.07 – 0.18 | **<0.001** | 0.12 | 0.07 – 0.17 | **<0.001** | 0.13 | 0.08 – 0.18 | **<0.001** |
| Time:WRTS | -0.00 | -0.01 – 0.01 | 0.832 | -0.00 | -0.01 – 0.01 | 0.804 | -0.00 | -0.01 – 0.01 | 0.697 | -0.00 | -0.01 – 0.00 | 0.578 |
| HCC |  |  |  |  |  |  |  |  |  | 0.14 | 0.01 – 0.27 | **0.036** |
| WRTS:HCC |  |  |  |  |  |  |  |  |  | 0.01 | 0.00 – 0.03 | **0.023** |
| Time:HCC |  |  |  |  |  |  |  |  |  | -0.02 | -0.05 – 0.00 | 0.095 |
| Time:WRTS:HCC |  |  |  |  |  |  |  |  |  | -0.00 | -0.00 – -0.00 | **0.042** |
| Marginal R<sup>2</sup> /<br>Conditional R<sup>2</sup> |  |  |  |  | 0.331 / 0.527 |  |  | 0.363 / 0.588 |  |  | 0.366 / 0.602 |  |

*Note.* TE = Trauma exposure, WRTS = Work-related trauma severity, HCC = Hair cortisol concentration, PCL = PTSD checklist for DSM-5, CI = Confidence interval.

The final model yielded a significant interaction between Time, WRTS, and HCC, which was further decomposed using simple slopes (see Table 3). Decomposing effects by Time and WRTS revealed a positive correlation between HCC and PTSD symptoms in firefighters with average and above-average WRTS (see Figure 1). No significant associations were evident at six or twelve months.

**Table 3.**
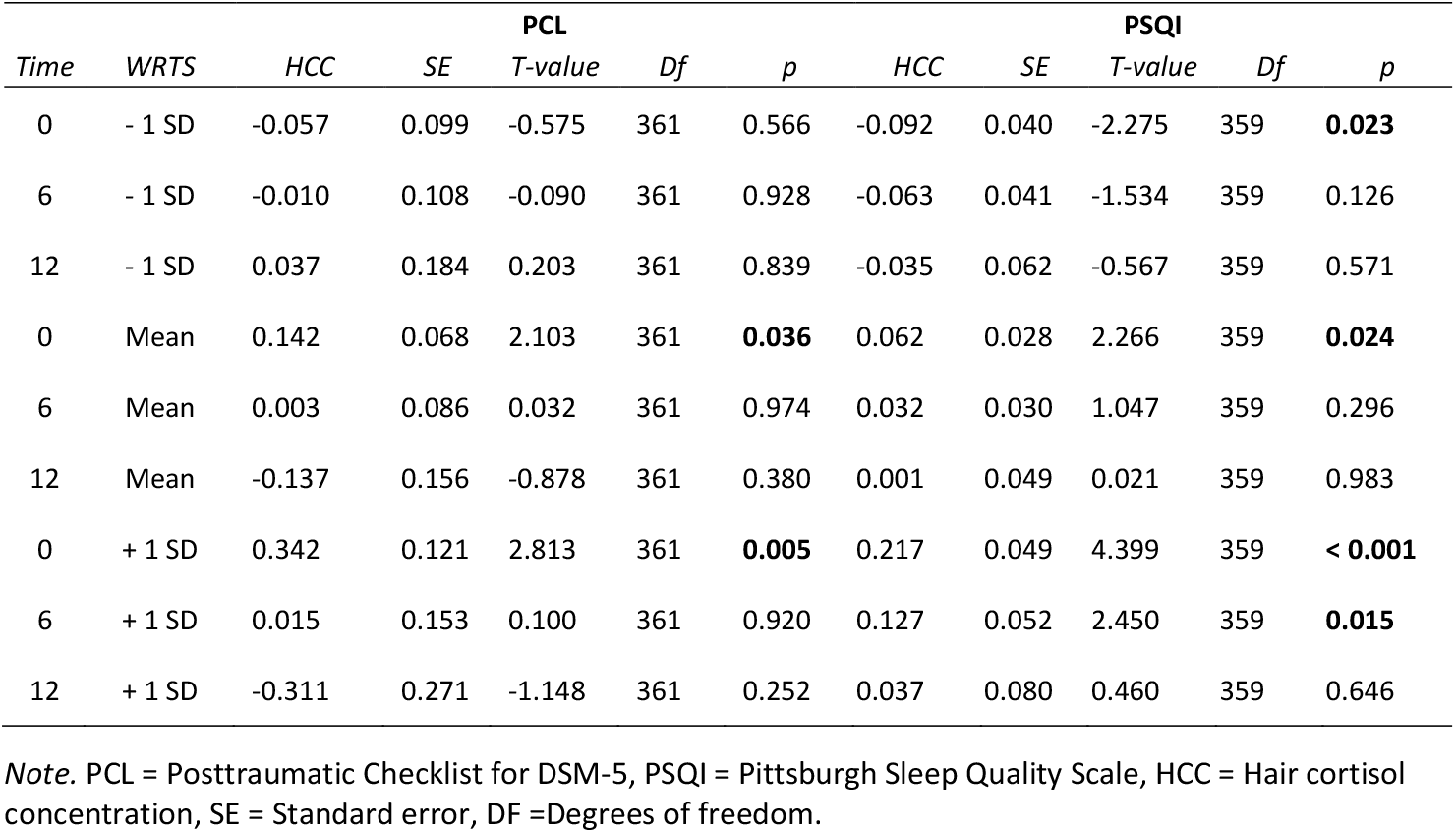
Simple slopes for different time points and levels of Work-related trauma severity (WRTS

**Figure 1.**
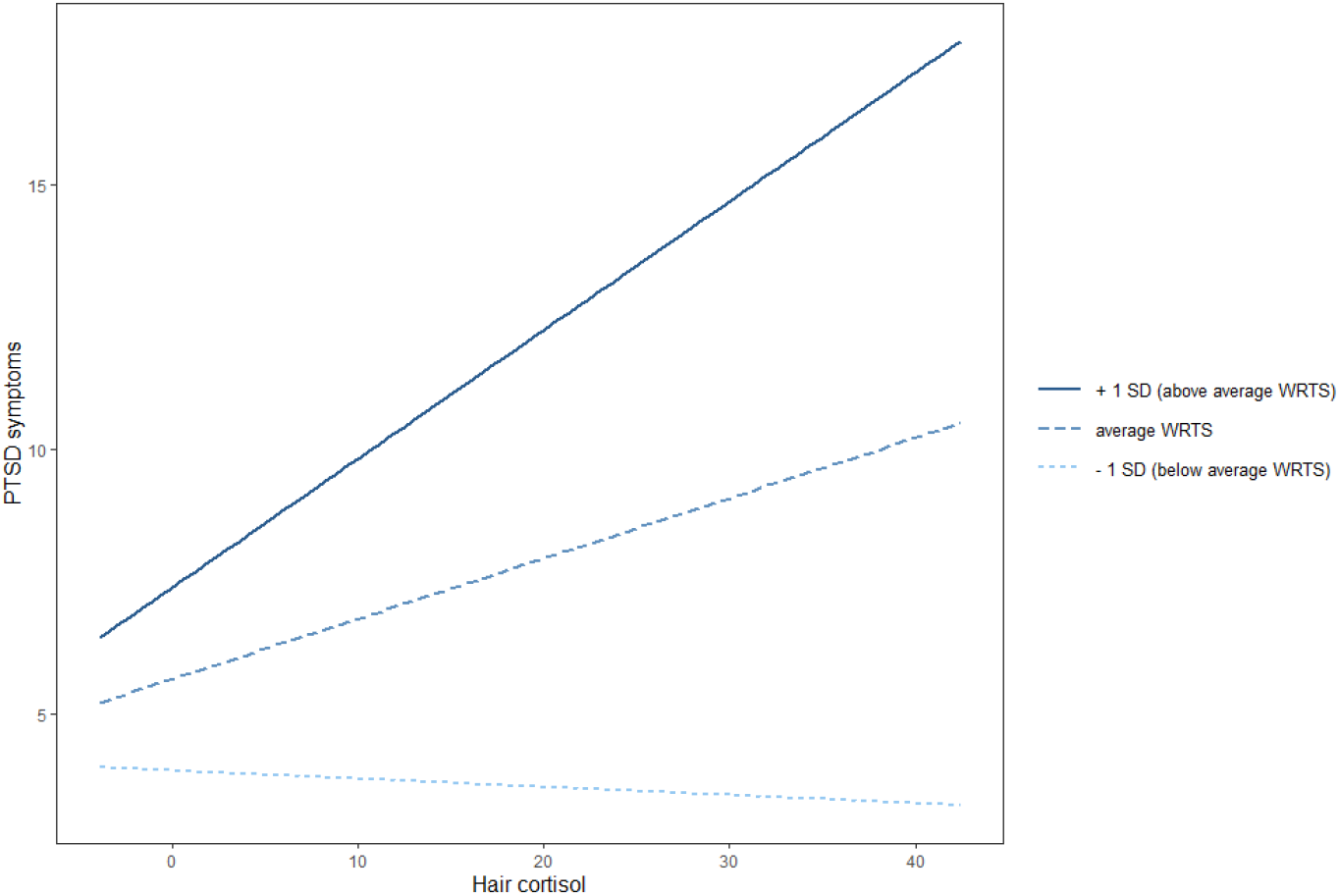
Association between Hair cortisol and PTSD symptoms for above average (+1 standard deviation), average (mean), and below average (−1 standard deviation) work-related trauma severity (WRTS) at T0 Note. SD = Standard deviation.

### Prediction of Sleep Disturbances Based on HCC

Repeated assessments of sleep disturbances were non-independent as reflected in an ICC of .628. As for PTSD symptoms, the model including a random intercept fitted our data significantly better than the baseline model (χ^2^_diff_ (1) = 122.10, *p* < .0001). However, including a random slope did not improve model fit (χ^2^_diff_ (2) = 0.14, *p* = .934), whereas including HCC at baseline resulted in further improvements (χ^2^_diff_ (4) = 21.13, *p* = .0003). Table 4 provides an overview of intercepts and slopes as well as the estimated variance accounted for by each model.

**Table 4.** Model summaries of linear mixed model analyses for sleep disturbances

| *Predictors* | Baseline model |  |  | Random intercept |  |  | Random intercept + Slope |  |  | Random intercept and Cortisol as predictor |  |  |
| --- | --- | --- | --- | --- | --- | --- | --- | --- | --- | --- | --- | --- |
|  | *Estimates* | *CI* | *p* | *Estimates* | *CI* | *p* | *Estimates* | *CI* | *p* | *Estimates* | *CI* | *p* |
| (Intercept) | 4.01 | 3.66 – 4.35 | **<0.001** | 4.07 | 3.66 – 4.48 | **<0.001** | 4.07 | 3.66 – 4.48 | **<0.001** | 4.10 | 3.70 – 4.50 | **<0.001** |
| Lifetime TE | 0.07 | 0.00 – 0.14 | **0.040** | 0.08 | -0.01 – 0.16 | 0.089 | 0.08 | -0.01 – 0.16 | 0.089 | 0.07 | -0.02 – 0.15 | 0.120 |
| Service years | -0.02 | -0.04 – -0.00 | **0.039** | -0.02 | -0.05 – 0.00 | 0.067 | -0.02 | -0.05 – 0.00 | 0.067 | -0.03 | -0.05 – -0.00 | **0.035** |
| Type of service | -0.09 | -0.28 – 0.09 | 0.310 | -0.09 | -0.32 – 0.14 | 0.461 | -0.09 | -0.32 – 0.14 | 0.462 | -0.08 | -0.31 – 0.14 | 0.466 |
| Psychopathology | 4.89 | 4.20 – 5.58 | **<0.001** | 5.15 | 4.29 – 6.00 | **<0.001** | 5.15 | 4.29 – 6.00 | **<0.001** | 4.88 | 4.04 – 5.73 | **<0.001** |
| Sex | -0.12 | -0.43 – 0.19 | 0.452 | -0.17 | -0.56 – 0.23 | 0.409 | -0.16 | -0.56 – 0.23 | 0.409 | -0.17 | -0.55 – 0.21 | 0.382 |
| Time | -0.01 | -0.05 – 0.02 | 0.418 | -0.01 | -0.04 – 0.02 | 0.460 | -0.01 | -0.04 – 0.02 | 0.459 | -0.01 | -0.04 – 0.01 | 0.367 |
| WRTS | 0.01 | -0.01 – 0.03 | 0.523 | 0.01 | -0.01 – 0.03 | 0.428 | 0.01 | -0.01 – 0.03 | 0.427 | 0.01 | -0.01 – 0.03 | 0.270 |
| Time:WRTS | -0.00 | -0.00 – 0.00 | 0.692 | -0.00 | -0.00 – 0.00 | 0.660 | -0.00 | -0.00 – 0.00 | 0.660 | -0.00 | -0.00 – 0.00 | 0.604 |
| HCC |  |  |  |  |  |  |  |  |  | 0.06 | 0.01 – 0.12 | **0.024** |
| WRTS:HCC |  |  |  |  |  |  |  |  |  | 0.01 | 0.01 – 0.02 | **<0.001** |
| Time:HCC |  |  |  |  |  |  |  |  |  | -0.01 | -0.01 – 0.00 | 0.228 |
| Time:WRTS:HCC |  |  |  |  |  |  |  |  |  | -0.00 | -0.00 – -0.00 | **0.030** |
| Marginal R<sup>2</sup> /<br>Conditional R<sup>2</sup> |  |  |  |  | 0.262 / 0.633 |  |  | 0.262 / 0.634 |  |  | 0.279 / 0.634 |  |

*Note.* TE = Trauma exposure, WRTS = Work-related trauma severity, HCC = Hair Cortisol Concentration, PCL = PTSD checklist for DSM-5, CI = Confidence interval.

The final model yielded a significant interaction between Time, WRTS, and HCC, which was further decomposed using simple slopes (see Table 3). Decomposing effects by Time and WRTS revealed a positive correlation between HCC and sleep disturbances in firefighters with average and above-average WRTS and a negative correlation between HCC and sleep disturbances in those with below-average WRTS at baseline (see Figure 2). After six months, firefighters with above-average WRTS still showed a positive correlation between baseline HCC and sleep disturbances (see Figure 3). No significant associations were evident at twelve months.

**Figure 2.**
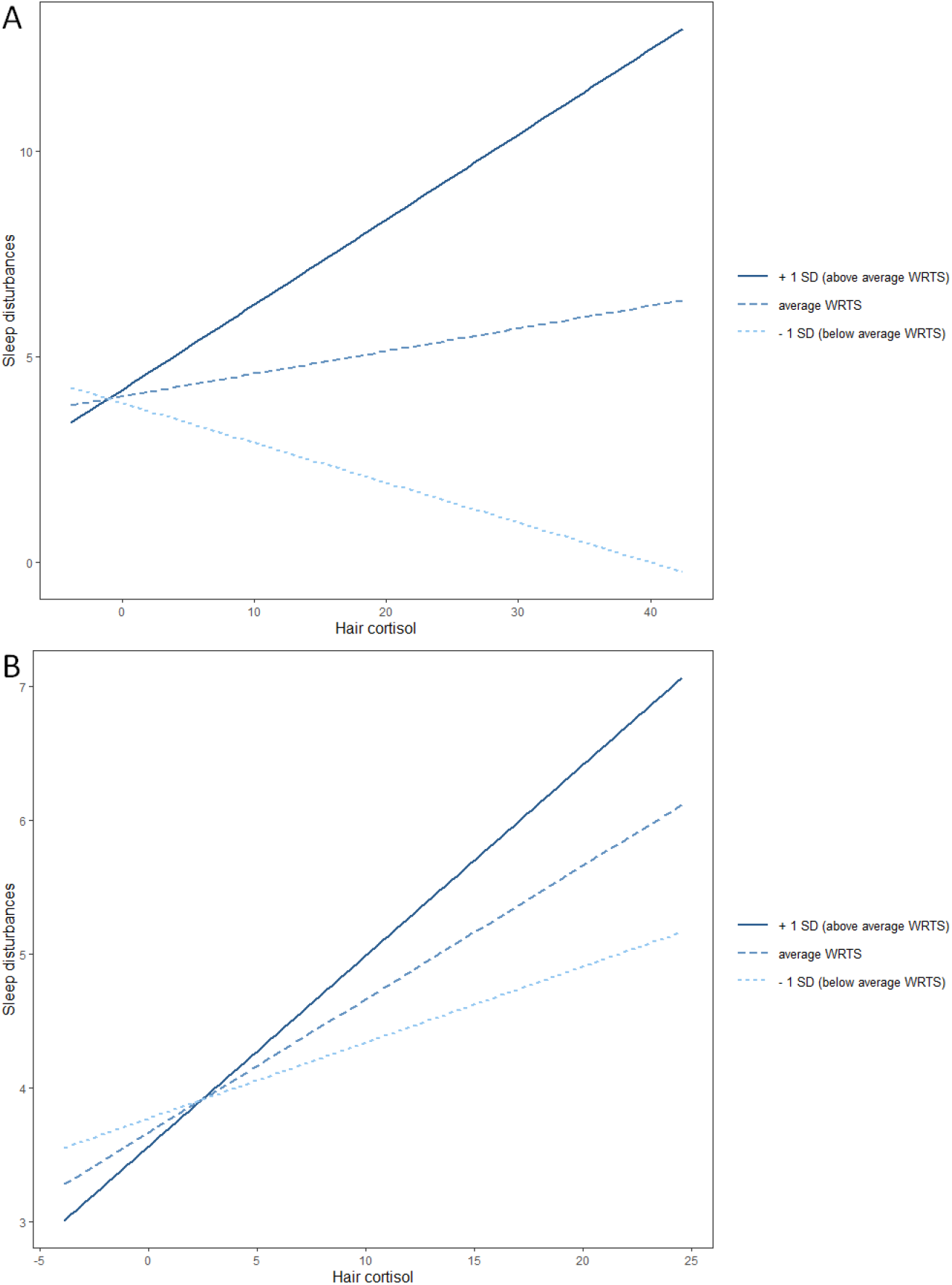
Association between Hair cortisol and Sleep disturbances for above average (+1 standard deviation), average (mean), and below average (−1 standard deviation) work-related trauma severity (WRTS) at T0 (A) and T1 (B) Note. SD = Standard deviation.

## Discussion

The current study aimed to investigate the association between HCC and PTSD symptoms and the temporal stability of this association. In addition, we examined whether associations are also evident for posttraumatic sleep disturbances. Analyses revealed that higher HCC was associated with higher baseline PTSD symptoms in firefighters with average and above-average WRTS. This pattern was no longer evident for symptoms measured after six and twelve months. Our analyses further revealed a positive association between HCC and baseline sleep disturbances in those with average and above-average WRTS and a negative association in those with below-average WRTS. After six months, those with above-average WRTS still showed a positive correlation between HCC and sleep disturbances. However, no association emerged after twelve months.

In contrast to our hypotheses, we found a positive rather than a negative association between HCC and PTSD symptoms. Though contradicting the notion that hyposecretion of cortisol is linked with PTSD symptoms in individuals with repeated traumatization, our findings align with some recent findings linking higher HCC with PTSD (Buchmüller et al., 2020; Pacella et al., 2017; Steudte et al., 2011; van den Heuvel et al., 2020). Two different explanations may account for these inconsistent findings. Firstly, it has been discussed that trauma timing may be a critical determinant of the direction of the association between cortisol and PTSD symptoms (Steudte-Schmiedgen et al., 2016). High cortisol levels as a short-term response and low cortisol levels as a long-term response to trauma exposure may thus similarly be linked to PTSD. In the current study, we did not assess trauma timing, since such an assessment is difficult to conduct in settings of repeated and ongoing trauma exposure. To further elucidate the time course of the association between HCC and PTSD, future prospective studies should thus aim to recruit trainee firefighters and use continuous ambulatory assessments to capture individual traumatic incidents and subsequent changes in HCC. Such a design would also strengthen causal interpretation of putative associations. Unfortunately, with 14 years of work experience on average, the majority of our sample had experienced several, heterogeneously spaced traumatic incidents prior to HCC assessment, limiting our ability to account for trauma timing and draw causal inferences.

Beyond trauma timing, van den Heuvel et al. (2020) recently suggested that patients with PTSD may show heightened cortisol levels in high-stress environments with ongoing stressors, whereas they may show reduced cortisol levels in low-stress environments. One could argue that the work environment of firefighters constitutes a high-stress environment with repeated stressors, which could account for the positive correlation that emerged between HCC and PTSD symptoms. The fact that only firefighters with average and above-average WRTS showed a positive association between cortisol levels and PTSD symptoms and that this correlation was stronger for those with above-average WRTS further aligns with the notion of subjective high-stress environments evoking a positive link between cortisol levels and PTSD symptoms. However, further research is needed to substantiate this claim.

Relatedly, extended work experience at baseline may have affected the direction of the association between HCC and PTSD symptoms in the current sample. Although we were not able to fully capture such effects in our design, we did find indications that extended work-related experiences was associated with higher WRTS and significantly contributed to the final model predicting sleep disturbances. In addition, assessing most firefighters after more than a decade of service may have introduced self-selection effects. That is, our sample likely consisted of firefighters who manage to maintain a high level of functioning across their career, whereas those strongly affected by work-related trauma likely terminate their service early on (Vargas de Barros et al., 2013). To address such effects, future studies should include subsamples of trainee firefighters and firefighters with substantial work experience. Such a design would allow to examine whether effects are robust across different career stages. Moreover, future studies in samples with advanced work experience should seek to explicitly investigate resilience as an alternative outcome of occupational trauma and potential associations with HCC. Previous findings suggest that HCC may be inversely correlated with resilience (van den Heuvel et al., 2020). However, this needs to be reinvestigated in the context of occupational trauma and in a longitudinal design comprising repeated assessments of resilience.

For posttraumatic sleep disturbances, we found highly similar associations as for PTSD symptoms: Those with average and above-average WRTS showed a positive correlation between cortisol levels and sleep disturbances. This finding is in line with previous studies showing an association between HCC and sleep disturbances in occupational settings (Wang et al., 2019; Zhang et al., 2020). Notably, our study is the first to demonstrate such an association in a sample of traumatized individuals. Beyond this positive association, we found a negative association between HCC and sleep disturbances in firefighters with below-average WRTS. Though unexpected, this finding could relate to the fact that we assessed posttraumatic sleep disturbances rather than sleep disturbances per se. As such, our results might be reflective of sleep disturbances comorbid with (sub-)clinical PTSD symptoms, which is supported by the high correlation between PCL and PSQI scores (see Table 1). The negative correlation between HCC and sleep disturbances may thus be interpreted within the framework of cortisol dysregulation in PTSD (Steudte-Schmiedgen et al., 2016) with some findings indicating that a negative association between HCC and symptoms might be specifically evident after mild trauma in low-stress environments (Steudte-Schmiedgen et al., 2015). In further support of this notion, our analysis of PTSD symptoms similarly revealed a negative, yet non-significant association between HCC and PTSD symptoms in those with below-average WRTS. However, caution is warranted in drawing strong conclusions and future research is needed to test whether trauma severity modulates the direction of the association between HCC and trauma symptoms.

Regarding the longitudinal course of the association between HCC and symptoms, we expected that associations would increase with time and subsequent trauma exposure. However, in contrast to this hypothesis, we only found a significant association between baseline cortisol and sleep disturbances after six months. No other associations emerged six or twelve months after baseline assessment. By suggesting that HCC may only be transiently related to PTSD symptoms, our results seem to contradict the notion that cortisol dysregulation is a predictor of PTSD symptoms. However, due to the high level of dropout, it is difficult to draw strong conclusions. Two alternative interpretations that may account for the current result pattern must thus be considered: Firstly, those with higher PTSD symptoms might have discontinued study participation after baseline, resulting in a lack of association at follow-up. To address this possibility, we conducted follow-up analyses comparing baseline PTSD levels of responders and non-responders at six and twelve months. Neither of these analyses revealed a significant difference (*p* > .05). However, further comparisons indicated that non-responders at six and twelve months had been in service for a fewer number of years (p < .05). Hence, systematic dropout of first responders with fewer service years may have brought about the lack of associations evident for symptoms measured at six and twelve months. No differences emerged for Sex, Type of service (volunteer vs. professional), Lifetime trauma exposure, Psychopathology, Baseline sleep quality, WRTS or Cortisol (*p* > .05).

Secondly, the substantial loss of participants might have brought about a loss of power, reducing the chance of detecting existing associations. Unfortunately, drop-out is difficult to prevent in occupational trauma settings as reflected in similarly high rates in previous studies (see e.g., Steudte-Schmiedgen et al., 2015). However, this issue can be partly addressed by collecting larger samples at baseline to ensure adequate power at subsequent follow-ups. Overall, in light of two former studies finding a longitudinal association between HCC and PTSD symptoms, two studies showing no such association, and our study similarly failing to do so, we believe that the current state of research is too sparse to draw any conclusions as to whether HCC may be a prospective correlate of PTSD symptoms. Future studies with large samples and prospective designs are thus needed to shed further light on this issue. Although interpretation of our findings is restricted with respect to PTSD symptoms, we did find that – for high WRTS – HCC predicted sleep disturbances 6 months later. This finding supports the notion that posttraumatic cortisol dysregulation is a precursor of sleep disturbances. In light of the high prevalence of posttraumatic sleep disturbances and their associated burden, these findings may be relevant for clinical practice and should be considered in the context of early prevention strategies. Moreover, previous studies discuss that cortisol dysregulation may facilitate sleep disturbances, which may in turn be a precursor of PTSD (Kobayashi & Delahanty, 2015; Otte et al., 2005). Hence, our findings may have indirect implications for PTSD development. Future research should address this further by modeling the complex associations between cortisol, sleep disturbances, and PTSD symptoms across time.

Beyond the limitations raised above, several other weaknesses of the current study design should be considered. Firstly, our sample consisted almost exclusively of male participants (92.72%). Since sex has been found to influence the association between cortisol and PTSD symptoms (Straub et al., 2017), this limits the generalizability of our findings to female populations. Secondly, to increase feasibility, our study design did not include a detailed assessment of which traumatic events happened between each time point and HCC was only sampled at baseline. We were therefore not able to model change in HCC after trauma and its influence on coping with subsequent trauma. Such investigations are needed to test the model proposed by Steudte-Schmiedgen et al. (2016). A prospective assessment approach would further enable measuring the exact number of traumatic events experienced, whereas the current study assessed different types of work-related traumatic incidents without accounting for the number of times that each of these incidents had occurred. In the same effort to increase feasibility, we did not include additional measures of cortisol such as salivary or serum cortisol levels. Such auxiliary measures are important to confirm that associations are evident across different assessment methods. Moreover, previous research indicates that HCC is influenced by ambient temperature (Boesch et al., 2015). Although firefighters use protective gear when exposed to high temperatures, future research should seek to include an additional measure of cortisol levels to rule out temperature-related effects.

Another limitation concerns the fact that we focused on occupational trauma. Although this allowed us to assess a homogenous group of participants, our findings might not generalize to trauma survivors from the general population. Relatedly, we included both professional and non-professional firefighters in our sample in order to reach the maximum attainable sample size. Although accounting for Type of service did not yield significant effects in our final models, future studies should consider assessing either professional or non-professional firefighters in order to exclude related sources of variance. Another limitation is that we did not collect baseline diagnoses of mental disorders in our sample (i.e., using structured clinical interviews). Moreover, we only assessed PTSD symptoms and sleep disturbances as continuous outcomes, which does not cover all possible outcomes of occupational trauma (e.g., depression, substance use disorders, burnout; Schäfer et al., 2020). Finally, although we controlled for lifetime trauma exposure, our assessment did not capture early life adversities and childhood trauma. Controlling for these variables and investigating their impact on the association between HCC and trauma symptoms should be considered by future research (see e.g., Schreier et al., 2015).

## Conclusions

To the best of our knowledge, this is the study with the largest sample to date investigating the prospective association between HCC and PTSD symptoms in traumatized adults. As such, it supports previous research in smaller samples showing that the direction of the association between HCC and PTSD symptoms may be positive under certain conditions (Buchmüller et al., 2020; Pacella et al., 2017). Our results further indicate that subjective trauma severity may be an important moderator of the direction of this association. Moreover, our study is the first that demonstrates a link between HCC and posttraumatic sleep disturbances. This link was also evident after six months, indicating that HCC has some predictive validity in the context of posttraumatic sleep disturbances. If consolidated by future research, these findings could have important clinical implications. That is, individuals with high cortisol levels could be identified early after trauma for strategic prevention. Such prevention should consider targeting sleep hygiene in order to prevent the development of posttraumatic sleep disorders.

## Data Availability

Data will be provided by the corresponding author upon reasonable request.

## Notes

### Competing Interest Statement

The authors have declared no competing interest.

### Funding Statement

This research did not receive any specific grant from funding agencies in the public, commercial, or not-for-profit sectors.

### Author Declarations

The study protocol was approved by the ethics committee of Behavioural and Social Sciences at the University of Groningen.

## References

Abbasi, M., Rajabi, M., Yazdi, Z., & Shafikhani, A. A. (2018). Factors affecting sleep quality in firefighters. Sleep and Hypnosis.

APA. (2013). Diagnostic and statistical manual of mental disorders (DSM-5®). American Psychiatric Pub.

Boesch, M., Sefidan, S., Annen, H., Ehlert, U., Roos, L., Van Uum, S., … & La Marca, R. (2015). Hair cortisol concentration is unaffected by basic military training, but related to sociodemographic and environmental factors. Stress, 18(1), 35–41.

Boeschoten, M., Bakker, A., Jongedijk, R., & Olff, M. (2014). PTSD checklist for the DSM-5 (PCL-5)– Nederlandstalige versie. Diemen: Arq Psychotrauma Expert Groep.

Born, J., Muth, S., & Fehm, H. L. (1988). The significance of sleep onset and slow wave sleep for nocturnal release of growth hormone (GH) and cortisol. Psychoneuroendocrinology, 13(3), 233–243.

Buchmüller, T., Lembcke, H., Busch, J., Kumsta, R., Wolf, O. T., & Leyendecker, B. (2020). Exploring hair steroid concentrations in asylum seekers, internally displaced refugees, and immigrants. Stress, 23(5), 538–545.

Buysse, D. J., Reynolds III, C. F., Monk, T. H., Hoch, C. C., Yeager, A. L., & Kupfer, D. J. (1991). Quantification of subjective sleep quality in healthy elderly men and women using the Pittsburgh Sleep Quality Index (PSQI). Sleep, 14(4), 331–338.

Carey, M. G., Al-Zaiti, S. S., Dean, G. E., Sessanna, L., & Finnell, D. S. (2011). Sleep problems, depression, substance use, social bonding, and quality of life in professional firefighters. Journal of occupational and environmental medicine/American College of Occupational and Environmental Medicine, 53(8), 928.

Colvonen, P. J., Straus, L. D., Stepnowsky, C., McCarthy, M. J., Goldstein, L. A., & Norman, S. B. (2018). Recent advancements in treating sleep disorders in co-occurring PTSD. Current Psychiatry Reports, 20(7), 1–13.

de Haart, R., Mouthaan, J., Vervliet, B., & Lommen, M. J. J. (2021). Avoidance learning as predictor of posttraumatic stress in firefighters. Behavioural Brain Research, 402, 113064.

Derogatis, L. R., & Melisaratos, N. (1983). The brief symptom inventory: an introductory report. Psychological Medicine, 13(3), 595–605.

Hughes, J. (2017). reghelper: Helper Functions for Regression Analysis. R package version 0.3.3. Retrieved from https://CRAN.R-project.org/package=reghelper.

Egberink, I. J. L., Harms, T., & Lommen, M. J. J. (2020). Psychometric properties of the Dutch revised sense of coherence scale in a firefighter sample. European Journal of Psychotraumatology, 11(1), 1759984.

Geronazzo-Alman, L., Eisenberg, R., Shen, S., Duarte, C. S., Musa, G. J., Wicks, J., Fan, B., Doan, T., Guffanti, G., & Bresnahan, M. (2017). Cumulative exposure to work-related traumatic events and current post-traumatic stress disorder in New York City’s first responders. Comprehensive Psychiatry, 74, 134–143.

Gulliver, S. B., Zimering, R. T., Knight, J., Morissette, S. B., Kamholz, B. W., Pennington, M. L., … & Meyer, E. C. (2021). A prospective study of firefighters’ PTSD and depression symptoms: The first 3 years of service. Psychological Trauma: Theory, Research, Practice, and Policy, 13(1), 44.

Jahnke, S. A., Poston, W. S. C., Haddock, C. K., & Murphy, B. (2016). Firefighting and mental health: Experiences of repeated exposure to trauma. Work, 53(4), 737–744.

Kessler, R. C., Aguilar-Gaxiola, S., Alonso, J., Benjet, C., Bromet, E. J., Cardoso, G., Degenhardt, L., de Girolamo, G., Dinolova, R. V., Ferry, F., Florescu, S., Gureje, O., Haro, J. M., Huang, Y., Karam, E. G., Kawakami, N., Lee, S., Lepine, J.-P., Levinson, D., … Koenen, K. C. (2017). Trauma and PTSD in the WHO World Mental Health Surveys. European Journal of Psychotraumatology, 8(sup5), 1353383.

Kim, J. I., Park, H., & Kim, J. H. (2018). The mediation effect of PTSD, perceived job stress and resilience on the relationship between trauma exposure and the development of depression and alcohol use problems in Korean firefighters: A cross-sectional study. Journal of Affective Disorders, 229, 450–455.

Kobayashi, I., & Delahanty, D. L. (2014). Awake/sleep cortisol levels and the development of posttraumatic stress disorder in injury patients with peritraumatic dissociation. Psychological Trauma: Theory, Research, Practice, and Policy, 6(5), 449.

Kreft, I. G., De Leeuw, J., & Aiken, L. S. (1995). The effect of different forms of centering in hierarchical linear models. Multivariate Behavioral Research, 30(1), 1–21.

Lewis-Schroeder, N. F., Kieran, K., Murphy, B. L., Wolff, J. D., Robinson, M. A., & Kaufman, M. L. (2018). Conceptualization, assessment, and treatment of traumatic stress in first responders: A review of critical issues. Harvard Review of Psychiatry, 26(4), 216.

Luo, H., Hu, X., Liu, X., Ma, X., Guo, W., Qiu, C., Wang, Y., Wang, Q., Zhang, X., & Zhang, W. (2012). Hair cortisol level as a biomarker for altered hypothalamic-pituitary-adrenal activity in female adolescents with posttraumatic stress disorder after the 2008 Wenchuan earthquake. Biological Psychiatry, 72(1), 65–69.

Meewisse, M.-L., Reitsma, J. B., De Vries, G.-J., Gersons, B. P., & Olff, M. (2007). Cortisol and post-traumatic stress disorder in adults: systematic review and meta-analysis. The British Journal of Psychiatry, 191(5), 387–392.

Mollayeva, T., Thurairajah, P., Burton, K., Mollayeva, S., Shapiro, C. M., & Colantonio, A. (2016). The Pittsburgh sleep quality index as a screening tool for sleep dysfunction in clinical and non-clinical samples: A systematic review and meta-analysis. Sleep Medicine Reviews, 25, 52–73.

Otte, C., Lenoci, M., Metzler, T., Yehuda, R., Marmar, C. R., & Neylan, T. C. (2005). Hypothalamic– pituitary–adrenal axis activity and sleep in posttraumatic stress disorder. Neuropsychopharmacology, 30(6), 1173–1180.

Pacella, M. L., Hruska, B., Steudte-Schmiedgen, S., George, R. L., & Delahanty, D. L. (2017). The utility of hair cortisol concentrations in the prediction of PTSD symptoms following traumatic physical injury. Soc Sci Med, 175, 228–234.

Pan, X., Kaminga, A. C., Wen, S. W., Wang, Z., Wu, X., & Liu, A. (2020). The 24-hour urinary cortisol in post-traumatic stress disorder: A meta-analysis. PLoS ONE, 15(1), e0227560.

Pan, X., Wang, Z., Wu, X., Wen, S. W., & Liu, A. (2018). Salivary cortisol in post-traumatic stress disorder: A systematic review and meta-analysis. BMC Psychiatry, 18(1), 324.

Petrowski, K., Wichmann, S., Pyrc, J., Steudte-Schmiedgen, S., & Kirschbaum, C. (2020). Hair cortisol predicts avoidance behavior and depressiveness after first-time and single-event trauma exposure in motor vehicle crash victims. Stress, 23(5), 567–576.

Pinheiro, J., Bates, D., DebRoy, S., Sarkar, D., Heisterkamp, S., Van Willigen, B., & Maintainer, R. (2017). Package ‘nlme’. Linear and nonlinear mixed effects models, version, 3(1).

Roberge, E. M., & Bryan, C. J. (2021). An integrated model of chronic trauma-induced insomnia. Clinical Psychology & Psychotherapy, 28(1), 79–92.

Rodenbeck, A., Huether, G., Rüther, E., & Hajak, G. (2002). Interactions between evening and nocturnal cortisol secretion and sleep parameters in patients with severe chronic primary insomnia. Neuroscience Letters, 324(2), 159–163.

Schäfer, S. K., Sopp, M. R., Staginnus, M., Lass-Hennemann, J., & Michael, T. (2020). Correlates of mental health in occupations at risk for traumatization: A cross-sectional study. BMC Psychiatry, 20(1), 1–14.

Schnell, T., Suhr, F., & Weierstall-Pust, R. (2020). Post-traumatic stress disorder in volunteer firefighters: influence of specific risk and protective factors. European Journal of Psychotraumatology, 11(1), 1764722.

Schreier, H. M., Enlow, M. B., Ritz, T., Gennings, C., & Wright, R. J. (2015). Childhood abuse is associated with increased hair cortisol levels among urban pregnant women. J Epidemiol Community Health, 69(12), 1169–1174.

Schumacher, S., Niemeyer, H., Engel, S., Cwik, J. C., Laufer, S., Klusmann, H., & Knaevelsrud, C. (2019). HPA axis regulation in posttraumatic stress disorder: A meta-analysis focusing on potential moderators. Neuroscience & Biobehavioral Reviews, 100, 35–57.

Singer, J. D., & Willett, J. B. (2005). Growth curve modeling. Encyclopedia of Statistics in Behavioral Science.

Soravia, L. M., Schwab, S., Walther, S., & Müller, T. (2021). Rescuers at risk: posttraumatic stress symptoms among police officers, fire fighters, ambulance personnel, and emergency and psychiatric nurses. Frontiers in psychiatry, 11, 1553.

Stalder, T., & Kirschbaum, C. (2012). Analysis of cortisol in hair–state of the art and future directions. Brain, Behavior, and Immunity, 26(7), 1019–1029.

Stalder, T., Steudte-Schmiedgen, S., Alexander, N., Klucken, T., Vater, A., Wichmann, S., Kirschbaum, C., & Miller, R. (2017). Stress-related and basic determinants of hair cortisol in humans: A meta-analysis. Psychoneuroendocrinology, 77, 261–274.

Steudte-Schmiedgen, S., Kirschbaum, C., Alexander, N., & Stalder, T. (2016). An integrative model linking traumatization, cortisol dysregulation and posttraumatic stress disorder: Insight from recent hair cortisol findings. Neuroscience & Biobehavioral Reviews, 69, 124–135.

Steudte-Schmiedgen, S., Stalder, T., Schönfeld, S., Wittchen, H.-U., Trautmann, S., Alexander, N., Miller, R., & Kirschbaum, C. (2015). Hair cortisol concentrations and cortisol stress reactivity predict PTSD symptom increase after trauma exposure during military deployment. Psychoneuroendocrinology, 59, 123–133.

Steudte, S., Kolassa, I.-T., Stalder, T., Pfeiffer, A., Kirschbaum, C., & Elbert, T. (2011). Increased cortisol concentrations in hair of severely traumatized Ugandan individuals with PTSD. Psychoneuroendocrinology, 36(8), 1193–1200.

Straub, J., Klaubert, L. M., Schmiedgen, S., Kirschbaum, C., & Goldbeck, L. (2017). Hair cortisol in relation to acute and post-traumatic stress symptoms in children and adolescents. Anxiety Stress Coping, 30(6), 661–670.

R Core Team (2013). R: A language and environment for statistical computing.

van den Heuvel, L. L., Acker, D., du Plessis, S., Stalder, T., Suliman, S., Thorne, M. Y., … & Seedat, S. (2020). Hair cortisol as a biomarker of stress and resilience in South African mixed ancestry females. Psychoneuroendocrinology, 113, 104543.

van den Heuvel, L. L., Stalder, T., du Plessis, S., Suliman, S., Kirschbaum, C., & Seedat, S. (2020). Hair cortisol levels in posttraumatic stress disorder and metabolic syndrome. Stress, 23(5), 577–589.

van Zuiden, M., Savas, M., Koch, S. B., Nawijn, L., Staufenbiel, S. M., Frijling, J. L., Veltman, D. J., van Rossum, E. F., & Olff, M. (2019). Associations among hair cortisol concentrations, posttraumatic stress disorder status, and amygdala reactivity to negative affective stimuli in female police officers. Journal of Traumatic Stress, 32(2), 238–248.

Vargas de Barros, V., Martins, L. F., Saitz, R., Bastos, R. R., & Ronzani, T. M. (2013). Mental health conditions, individual and job characteristics and sleep disturbances among firefighters. Journal of Health Psychology, 18(3), 350–358.

Wang, C., Dai, J. M., & Li, J. (2019). Mediating effects of hair cortisol on the mutual association of job burnout and insomnia: A retrospective exploratory study. J Psychiatr Res, 117, 62–67.

Werner, G. G., Riemann, D., & Ehring, T. (2020). Fear of sleep and trauma-induced insomnia: a review and conceptual model. Sleep Medicine Reviews, 101383.

Wortmann, J. H., Jordan, A. H., Weathers, F. W., Resick, P. A., Dondanville, K. A., Hall-Clark, B., Foa, E. B., Young-McCaughan, S., Yarvis, J. S., & Hembree, E. A. (2016). Psychometric analysis of the PTSD Checklist-5 (PCL-5) among treatment-seeking military service members. Psychological Assessment, 28(11), 1392.

Zhang, Y., Shen, J. Y., Zhou, Z. Q., Sang, L. L., Zhuang, X., Chu, M. J., Tian, T., Xiao, J., & Lian, Y. L. (2020). Relationships among shift work, hair cortisol concentration and sleep disorders: A cross-sectional study in China. Bmj Open, 10(11), e038786.

